# Prefrontal cortex connectivity profiles distinguish rapid from slow responders to deep brain stimulation in obsessive-compulsive disorder

**DOI:** 10.64898/2026.05.26.26353680

**Authors:** Sarah S. Soubra, Arnav Garyali, Reem El Jammal, Jonathan H. Bentley, Thomas A. Hamre, Nisha Giridharan, Corey P. St. Romain, Kasra A. Mansourian, Katherine E. Kabotyanski, Grace T. Nitcheu, Vinayak M. Belavadi, Melissa Ryan, Hideo Suzuki, Nora Vanegas-Arroyave, Melissa Franch, Eleonora Bartoli, Eric A Storch, Garrett P. Banks, Wayne K. Goodman, Nicole R. Provenza, Sameer A. Sheth, Sarah R. Heilbronner

## Abstract

**Introduction:** Deep brain stimulation (DBS) of the ventral capsule/ventral striatum (VC/VS) can benefit patients with treatment-refractory obsessive-compulsive disorder (OCD). However, time to respond post-operatively ranges from weeks to over a year. We examined neuroanatomical determinants of this variability.

**Methods:** We studied 16 treatment-refractory OCD patients who responded to VC/VS DBS, classifying them as rapid (≤3 months) or slow (>3 months) responders. We compared contact locations along anterior-posterior, dorsal-ventral, and medial-lateral axes. In 11 patients with diffusion-weighted magnetic resonance imaging (dMRI), we utilized volumes of tissue activated (VTAs) for both initial and most recent effective DBS settings to filter tractograms of the anterior limb of the internal capsule to 11 predefined prefrontal cortical regions. We analyzed streamline counts as a proxy for connectivity strength with mixed-effects models.

**Results:** Rapid (n=8) and slow (n=8) responders exhibited a clear bimodal distribution of time-to-response, supported by a Bayesian Information Criterion difference (Δ BIC) of 9.14. Rapid responders’ right-hemisphere contacts were positioned more superiorly, and there was a trend toward their left-hemisphere contacts being positioned more posteriorly. Connectivity fingerprints and mixed-effects modeling showed greater dorsolateral prefrontal cortex engagement in rapid responders than in slow responders, whereas slow responders showed enhanced central orbitofrontal cortex connectivity over time.

**Discussion:** Variability in VC/VS contact placement corresponds to distinct prefrontal cortical connectivity patterns and response timelines. Patient-specific targeting and connectivity-informed programming may accelerate response to treatment.

## 1. Introduction

Obsessive-compulsive disorder (OCD) is a chronic and debilitating psychiatric condition characterized by intrusive, anxiety-evoking thoughts leading to repetitive mental or behavioral activities to relieve the associated distress (Storch et al., 2008). While some patients achieve symptom control through pharmacologic and/or behavioral therapies, approximately 10% of patients do not, necessitating alternative approaches (Hirschtritt et al., 2017; Raviv et al., 2020). For these individuals, deep brain stimulation (DBS) is a promising neuromodulatory intervention. Current DBS targets include regions in and around the anterior limb of the internal capsule (ALIC), such as the ventral capsule/ventral striatum (VC/VS), ventral capsule/bed nucleus of the stria terminalis (VC/BNST), nucleus accumbens (NAc), subthalamic nucleus (STN), and inferior thalamic peduncle (Greenberg et al., 2006; Mallet et al., 2008a; Goodman et al., 2010; Luyten et al., 2016; Raviv et al., 2020; Provenza et al., 2021, 2024; Sheth et al., 2026). Approximately 66% of OCD DBS patients exhibit a clinical response at two years follow-up, with response defined as a ≥35% decrease in the Yale-Brown Obsessive Compulsive Scale (Y-BOCS).

Although DBS shows promise in alleviating OCD symptoms amount treatment refractory patients, clinical response trajectories are variable, suggesting that individual differences in anatomy or electrode placement may play a role. A subset of patients exhibits rapid clinical improvement within weeks of stimulation activation. For example, in one case study, the patient experienced partial remission within four weeks of DBS (Park et al., 2019). In another cohort, half of patients achieved a Y-BOCS reduction of >50% by month three (Mallet et al., 2008b). Others, however, may require several months to a year to achieve even modest benefit. In one study, while 28% of patients met responder criteria at one month, the response rate increased to >60% at two years (Greenberg et al., 2010). The heterogeneity in time to response underscores the need to better understand population statistics of each cohort and neuroanatomical substrates of differential trajectories of response.

One possibility is that response time variability may reflect differences in stimulation location. Early applications of DBS for OCD focused on targeting more anterior structures in the ALIC, mirroring the lesion site used in the anterior capsulotomy (Rauch, 2003). Over time, the DBS target has shifted more posteriorly in the ALIC, toward the BNST (Menchón et al., 2021; Naesström et al., 2021). A systematic review showed that anteriorly directed DBS results in ∼20% responders at 6 months and ∼36% at long-term follow-up, whereas for posteriorly directed DBS these numbers are ∼50% and ∼66% (Acevedo et al., 2021). One possibility is that posteriorly placed leads modulate a denser convergence of fibers from functionally distinct regions (Baldermann et al., 2019). Engagement of these denser white matter tracts thus offers an opportunity to modulate more distributed networks rather than isolated regions.

Indeed, the ALIC is topographically organized, with fibers from different prefrontal cortical (PFC) regions occupying distinct but overlapping portions of the bundle (Lehman et al., 2011; Safadi et al., 2018). Despite an increasingly rich literature on the particular white matter bundles associated with response (Hartmann et al., 2015; Coenen et al., 2017; Baldermann et al., 2019; Li et al., 2020; Graat et al., 2022; Widge et al., 2022; Gadot et al., 2024), most studies have focused on response magnitude or presence, with limited attention to the temporal course of symptom improvement. This gap represents a critical barrier to optimizing DBS targeting and programming optimization in a personalized, time-sensitive manner.

Here, we first evaluate response latencies across a cohort (N=16) of OCD patients who underwent DBS. Second, we assess the stimulation target locations for rapid and slow responders. Finally, we leverage patient-specific diffusion-weighted magnetic resonance imaging (dMRI) tractography to test whether connectivity between stimulation sites and specific prefrontal subregions distinguishes rapid from slow responders. By integrating tractography with response latency, this study aims to provide a more personalized, temporally sensitive approach to DBS targeting, one that considers not only where to stimulate, but also how cortical network engagement profiles might influence the speed of clinical response.

## 2. Methods

### Participants

We retrospectively analyzed a cohort of 16 patients with OCD treated with DBS targeting the VC/VS or VC/BNST who were classified as clinical responders. Clinical response was defined as a ≥ 35% reduction in Y-BOCS score from the baseline assessment preceding DBS activation (Tolin et al., 2005). Based on the observed response latencies (see Results), we dichotomized patients into fast responders, who met response criteria within three months of DBS activation, and slow responders, who required more than three months to respond to treatment. Non-responders were excluded from analysis. Data included in this paper are part of two clinical trials (NCT04806516 and NCT05915741 on clinicaltrials.gov).

### MRI Acquisition

We obtained high-resolution anatomical and diffusion-weighted MRI scans on a 3 Tesla Siemens MAGNETOM Prisma Fit scanner (Siemens Healthineers, Erlangen, Germany) with a 32-channel head coil at the Baylor College of Medicine (BCM) Core for Advanced Magnetic Resonance Imaging (CAMRI).

In the NCT04806516 study, we acquired T1-weighted Magnetization Prepared Rapid Gradient Echo (MPRAGE) sequence with 256 sagittal volumes, voxel size = 0.8 mm isotropic, TR = 2400 ms, TE = 2.24 ms, flip angle = 8° and FOV = 256 × 282 × 205 mm (except for one subject whose MPRAGE was acquired with TE = 2.41 ms and FOV = 224 × 224 × 179 mm); T2-weighted Sampling Perfection with Application-optimized Contrasts using different flip angle Evolution (SPACE) sequence with 208 sagittal volumes, voxel size = 0.8 mm isotropic, TR = 3200 ms, TE = 565 ms, flip angle = 120° and FOV = 240 × 256 × 166 mm (except for one subject whose SPACE was acquired with 256 sagittal volumes, voxel size = 0.7 mm isotropic and FOV = 224 × 224 × 179 mm); and multi-shell multi-band diffusion-weighted MRI sequences with 92 axial slices × 99 volumes (including 7 volumes of b = 0 s/mm^2^ and 46 directions each for b = 1250 and 2500 s/mm^2^), voxel size = 1.5 mm isotropic, TR = 3200 ms, TE = 87 ms, flip angle = 90°, FOV = 210 × 210 × 138 mm, pixel bandwidth = 1700 Hz and imaging frequency = 123.25 MHz (two sets of images acquired with opposite phase encoding direction).

In the NCT05915741 study, we acquired T1-weighted MPRAGE sequence with 256 sagittal volumes, voxel size = 0.7 mm isotropic, TR= 2400ms, TE = 2.41 ms, flip angle = 8° and FOV = 224 × 224 × 179 mm; T2-weighted SPACE sequence with 208 sagittal volumes, voxel size = 0.8 mm isotropic, TR = 3200 ms, TE = 563 ms, flip angle = 120° and FOV = 240 × 256 × 166 mm; and multi-band high-angular-resolution diffusion-weighted MRI sequences with 92 axial slices x 268 volumes using an anterior-posterior phase encoding direction (including 12 volumes of b = 0 s/mm^2^ and 256 diffusion-weighted volumes at b = 1000 s/mm^2^) and 92 axial slices x 7 volumes of b = 0 s/mm^2^ using a posterior-anterior phase encoding direction, voxel size = 2.0 mm isotropic, TR = 4230 ms, TE = 65 ms, flip angle = 90°, FOV = 256 × 256 × 128 mm, pixel bandwidth = 2300 Hz and imaging frequency ≈ 123.25 MHz.

Additionally, we obtained postoperative non-contrasted helical CT scans, performed following implantation, extending from skull base to vertex for lead localization.

### DBS Programming

DBS was activated 25.4 ± 16.9 (range: 7-73) days after lead implantation. The time to initiation of therapy differed across patients due to clinical considerations and scheduling constraints. We optimized stimulation through serial programming sessions using standard clinical assessments and self-reporting of energy, mood, and anxiety, as well as changes in OCD severity as measured by the Y-BOCS between visits (Goodman et al., 2020). We recorded stimulation parameters, including amplitude, frequency, and pulse width, and active DBS contacts at each programming visit. Following identification of clinical responders, we extracted the active contact(s) and stimulation settings from (i) the initial programming visit and (ii) the visit immediately preceding clinical response, representing the earliest stimulation configuration that led to clinically meaningful improvement.

We defined response latency as the interval, in days, between the initial programming session and the first visit at which the patient met clinical response criterion, with a ≥35% reduction in Y-BOCS relative to the post-operative, pre-programming baseline.

### Lead Localization

To explore the relationship between electrode placement and response latency, we localized the contacts activated at clinical response in Montreal Neurological Institute (MNI) space using Lead-DBS v3.0 (Neudorfer et al., 2023). We began with a linear co-registration of the patient’s pre-operative MRI and post-operative CT with Advanced Normalized Tools (Avants et al., 2008). We then normalized the co-registered images to the MNI non-linear 2009b space using ANT’s Symmetric Normalization (SyN) algorithm. We brainshift corrected using the coarse mask tool to account for post-operative linear deformation of brain tissue (Schönecker et al., 2009). Following co-registration, the Precise and Convenient Electrode Reconstruction for Deep Brain Stimulation (PaCER) toolbox automatically pre-reconstructed electrode trajectories and we performed manual refinements as needed (Husch et al., 2018). After reconstruction, we exported the MNI152 coordinates of active contacts at clinical response to compare rapid and slow responder cohorts in the medial-lateral, anterior-posterior, and superior-inferior axes in MNI152 space.

### Tractography

To probe structural connectivity differences underlying variable response latency, we analyzed dMRI in 11 of 16 responders with available dMRI data (n=11; 5 rapid responders, 6 slow responders). Five patients were excluded: two lacked diffusion scans, two had undergone acquisition protocols that were inconsistent with the remainder of the cohort, and one had undergone a pre-DBS capsulotomy, introducing a potential confound for ALIC tractography. MRtrix3 (v3.0) preprocessed dMRI data (Tournier et al., 2019), incorporating denoising, Gibbs ringing removal, FSL’s motion and eddy current correction, and ANT’s bias field correction. Constrained Spherical Deconvolution (CSD) estimated fiber orientation distributions (FOD) in MRtrix3.

We used the Anatomically Constrained Tractography (ACT) framework for probabilistic tractography. Within the ACT framework, we adjusted the 5-tissue-type mask to allow streamlines to pass through the caudate, putamen, and NAc, as many ventral PFC–ALIC fibers course through fascicles embedded in these subcortical structures (Lehman et al., 2011). To generate 1 million streamlines traversing the ALIC, we created the ALIC mask from the Johns Hopkins University ICBM-DTI-81 White Matter Labeled Atlas (Mori et al., 2008). In addition, we extracted 11 PFC subregions per hemisphere using the FreeSurfer’s Desikan-Killiany atlas (Desikan et al., 2006). These regions included the caudal middle frontal (CMF), rostral middle frontal (RMF), superior frontal (SF), caudal anterior cingulate (cACC), rostral anterior cingulate (rACC), pars opercularis, pars triangularis, par orbitalis, central orbitofrontal (cOF), and medial orbitofrontal (mOF). We divided the rACC region of interest (ROI) into dorsal (drACC) and ventral (vrACC) components at the level of the subcallosal cingulate cortex (z = +4 mm in MNI152 space) due to the unique trajectory of ventral fibers relative to dorsal fibers (Sretavan et al., 2024; El Jammal et al., 2026). All ROIs were registered to each subject’s native space using ANTs. We then performed probabilistic tractography of the ALIC using the MRtrix3 iFOD2 algorithm, with one million tracks generated per hemisphere from the ALIC seed. Using the “tckedit” function, we extracted streamline subsets passing through the ALIC and terminating in each PFC ROI.

After manually constructing fibers traversing the ALIC to the 11 predefined PFC regions, we utilized subject-specific volumes of tissue activated (VTA) generated using the Simbio FieldTrip Toolbox (Horn et al., 2017) in LeadDBS as inclusion filters to quantify the streamlines passing through the activated tissue and terminating in each PFC region. We modeled VTA masks using the stimulation parameters recorded at each patient’s first programming session and at the time of clinical response, yielding per-timepoint, per-hemisphere counts to each PFC ROI.

To summarize projection patterns by response latency and treatment time point, we generated per-hemisphere “connectivity fingerprint” plots in Python (v3.12.2) depicting the distribution of VTA-traversing streamlines across PFC regions over the treatment course.

### Statistical Analysis

We assessed the distribution of days to response in our patient cohort to determine whether response latency reflected two distinct distributions, indicating evidence of bimodality. To do so, we calculated the change in Bayesian Information Criterion (ΔBIC) to test for single versus two-component Gaussian fits and Asherman’s D to evaluate the degree of separation between the two distributions.

Following lead localization, we compared the spatial distribution of active contacts at response in slow and rapid responders along the medial-lateral (x) axis, anterior-posterior (y) axis, and superior-inferior (z) axes using false discovery rate (FDR)-corrected permutation tests.

We tested whether the variability of PFC connectivity differs by response latency and treatment timepoint using a linear mixed-effects model to predict raw streamline counts. Fixed effects were response type (rapid vs. slow), treatment timepoint from which each respective VTA was generated (initial programming visit vs. programming visit immediately preceding response), hemisphere (left or right), and region (one of 11 predefined PFC regions). Participant ID was included as a mixed effect with a random intercept to account for repeated measures within subjects. Post-hoc analysis included the assessment of interactions between response latency, treatment timepoint, and region, using the FDR to correct for multiple comparisons.

## Results

### Clinical Outcomes and Response Latency

Of the 16 patients classified as responders, we identified two response latency profiles. Eight patients (50%) were clinical responders within three months of their initial programming session with a mean response latency of 32.8 ± 18.8 days (range: 13-65 days), whereas the remaining eight (50%) were slow responders with a mean response latency of 293.5 ± 131.4 days (range: 154-489 days). Testing supported bimodality, with a two-component Gaussian mixture favored over a single Gaussian by 9.14 BIC points and well-separated curves (Asherman’s D = 2.66) (Figure 1A). Nonresponders were excluded from our analyses. Demographic data, including sex (p =1) and age (p = 0.29), OCD illness duration in years (p = 0.26), and baseline OCD score (p = 0.09) did not significantly differ between the two cohorts. Moreover, long-term follow-up scores, taken at the assessment nearest to the two-year mark, showed no significant group difference (p = 0.14).

**Figure 1.**
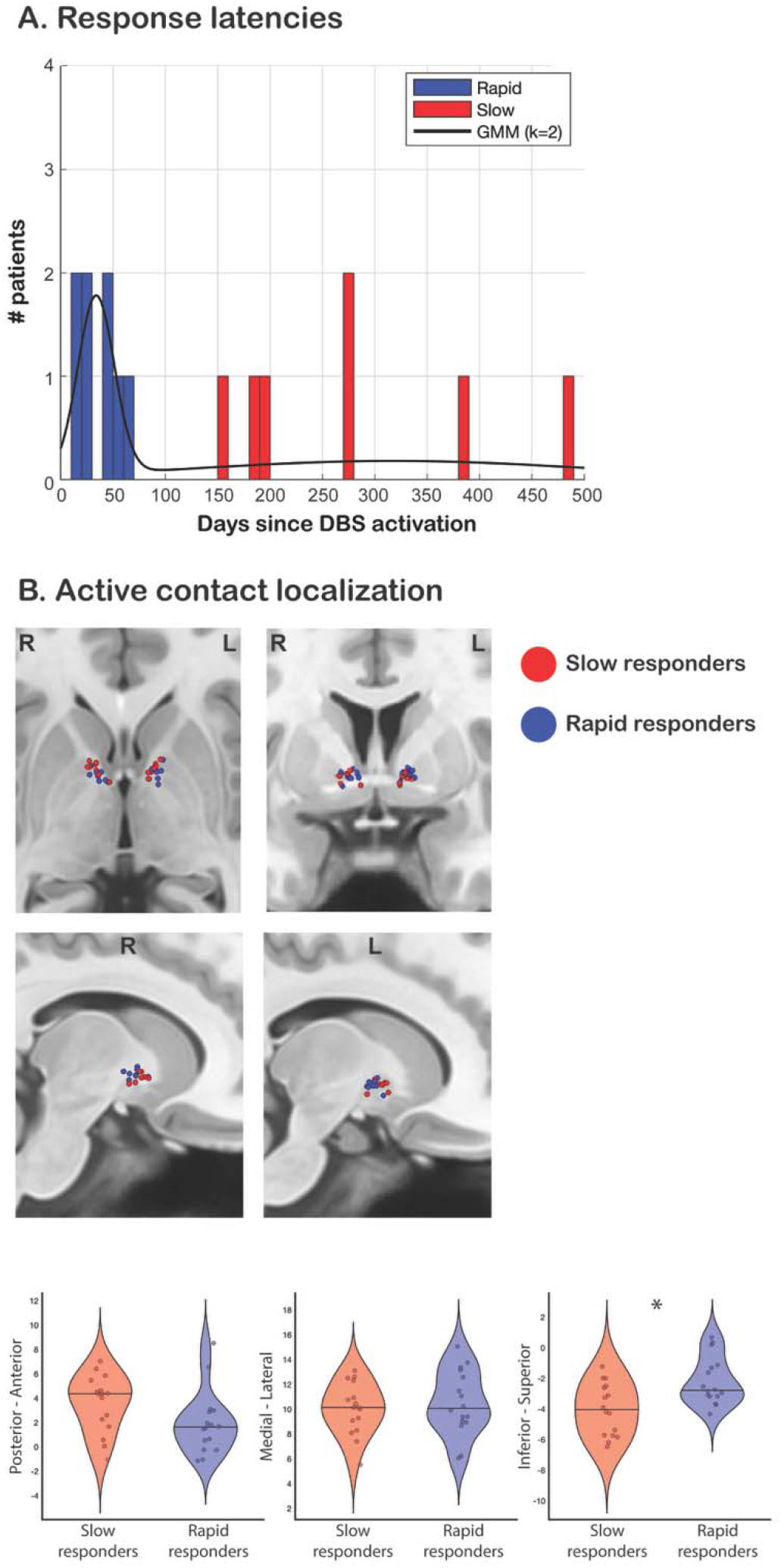
Time to response is bimodal and linked with specific lead locations. A. Time to response has a bimodal distribution. B. Localization of active contacts shows overlapping, but different, placement for rapid vs slow responders. Rapid responders are placed significantly superior relative to slow responders (p = 0.01) (bottom, right); other axes (bottom, left and middle) are not significant.

### Active Contact Localization

Active-contact maps in MNI space revealed cohort-specific trends in anatomic location. Comparing MNI coordinates of active contacts across axes by hemisphere, rapid responders exhibited more superiorly-positioned active contacts (*p* = 0.01) and trended to a more posterior location than slow responders (*p* = 0.1). On axial views, these contacts appeared to more often lie posterior to the anterior commissure (Figure 1B). No group differences were observed in the medial-lateral axes. All electrodes’ MNI coordinates are found in Supplemental Table 1.

### Structural Connectivity to the Prefrontal Cortex

We performed subject-specific tractography in 11 clinical responders (five rapid, six slow) with available dMRI. For each subject, we reconstructed tracts through the ALIC to 11 PFC regions in each hemisphere. We quantified streamlines that intersected VTAs defined at the initial programming session and at the response timepoint. Figure 2 shows fingerprint plots summarizing these distributions stratified by hemisphere, response latency, and treatment timepoint. Qualitative inspection reveals distinct connectivity patterns between groups and across timepoints.

**Figure 2.**
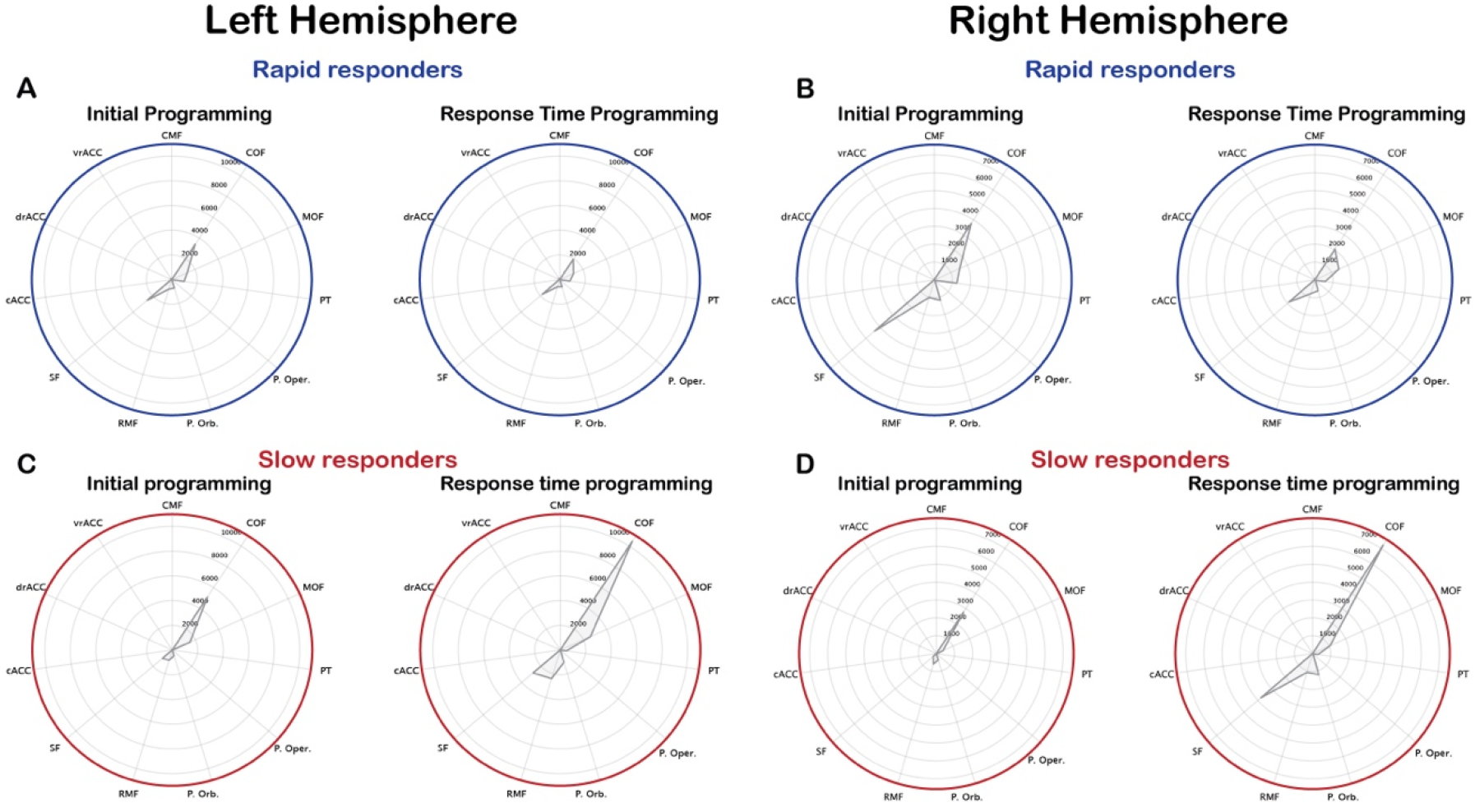
PFC connectivity profiles for rapid and slow responders at initial and response status programming dates. Amplitude indicates streamline count. A,B Streamline connectivity for rapid responders at initial programming in the left (A, left) and right (B, left) hemispheres show strong engagement with the superior frontal cortex (SF), particularly in the right hemisphere. Connectivity slightly declines at treatment timepoint immediately preceding response in the left (A, right) and right (B, right) hemispheres due to changes in stimulation parameters in several rapid responders. C,D Streamline connectivity for slow responders at initial programming in the left (C, left) and right (D, left) hemispheres show minimal engagement of the SF, but substantial engagement of the central orbitofrontal cortex (cOF). COF and SF engagement increase at the response timepoint in the left (C, right) and right (D, right) hemispheres. cACC=caudal anterior cingulate cortex; CMF=caudal middle frontal cortex; drACC=dorsal rostral anterior cingulate cortex; cOF=central orbitofrontal cortex; mOF=medial orbitofrontal cortex; P. Oper.=pars opercularis; P. Orb.=pars orbitalis; P. Tr.=pars triangularis; RMF=rostral middle frontal; SF=superior frontal; vrACC=ventral rostral anterior cingulate cortex.

Qualitative evaluation of connectivity fingerprints revealed different patterns over time for rapid and slow responders (Figure 2). Notably, VTA sizes at the initial programming session did not significantly differ between the cohorts (left hemisphere: *p* = 0.86; right hemisphere: *p* = 0.42), indicating that differences in initial network engagement were likely attributable to lead positioning rather than initial stimulation volume. In the left hemisphere, rapid responders mainly showed engagement of the cOF and SF cortex, with network engagement reducing over treatment time (Figure 2A). In the right hemisphere, there was a similar pattern for rapid responders, but with more streamlines engaged overall (Figure 2B). Slow responders had a different pace of network engagement over time. They exhibited an increase in overall streamline counts bilaterally from the initial programming to response in both hemispheres, most notably within the SF and cOF cortex (Figure 2C,D).

Indeed, mixed-effects linear modeling corroborated these qualitative observations (Figure 3). Across patients, engagement of the cOF, SF, RMF, and mOF were strong determinants of overall raw streamline counts, showing the largest and most significant positive associations with streamline counts and suggesting strong engagement of these regions (p < 0.001 in cOF and SF, p < 0.01 in mOF, and p < 0.05 in RMF) in all responders regardless of response latency. A significant *Treatment Time x Responder Type* interaction (*p*=0.001) indicated that, compared to rapid responders, slow responders exhibited a greater increase in total streamline counts at the response time point relative to the initial time point (Figure 3A).

**Figure 3.**
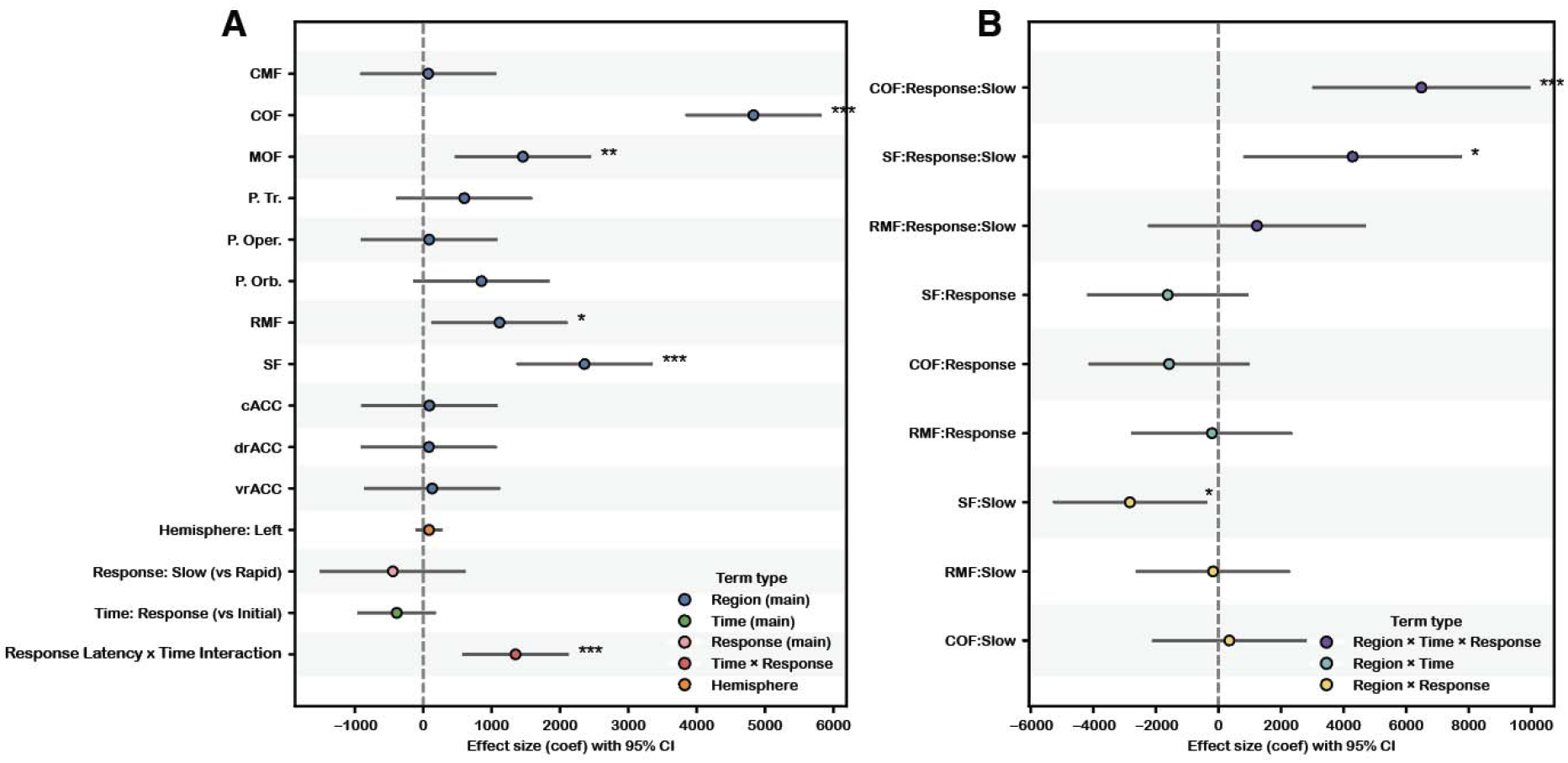
Results of mixed-effects linear modeling. A. Significant regional determinants of streamline counts revealed significant contributions from central orbitofrontal (cOF), medial orbitofrontal, rostral middle frontal, and superior frontal (SF) cortices. A Responder Type (rapid versus slow) × Treatment Time (initial versus response) interaction showed an increase in the number of streamlines captured over time in slow responders, but a decrease in rapid responders. B. Adding regional interaction terms showed significant interactions with the cOF and SF. Slow responders show increased streamline representation in the cOF cortex throughout the treatment course and lower SF streamline representation relative to rapid responders, but this representation increases over time. cACC=caudal anterior cingulate cortex; CMF=caudal middle frontal cortex; drACC=dorsal rostral anterior cingulate cortex; cOF=central orbitofrontal cortex; mOF=medial orbitofrontal cortex; P. Oper.=pars opercularis; P. Orb.=pars orbitalis; P. Tr.=pars triangularis; RMF=rostral middle frontal; SF=superior frontal; vrACC=ventral rostral anterior cingulate cortex. (only a subset of results shown; full results in the Supplement) * = p<0.05; ** = p<0.01; *** = p<0.001

Since the increase in streamlines in the slow responder group may be responsible for the improvement of symptoms, we conducted region-specific post-hoc tests to determine whether these changes were localized to specific frontal regions (Figure 3B). Adding the 11 frontal regions as an interaction variable revealed that at baseline, slow responders had significantly fewer SF streamlines (*p* = 0.02) compared to rapid responders, but exhibited a significantly greater increase in SF cortical recruitment from initial programming to response (*p* = 0.02), consistent with the idea that progressive SF cortical recruitment may have contributed to eventual response in slow responders. This is particularly pertinent in the right hemisphere (Figure 2B,D) For the cOF, slow responders demonstrated a statistically significant increase in streamline counts relative to rapid responders (*p* = 0.0003), though baseline cOF counts did not differ between groups (*p* = 0.78). No other regions exhibited significant variability. Overall, these findings suggest that the SF cortex could play a more central role in determining time-to-response than cOF recruitment, which increased over treatment time but did not initially differ between response cohorts. Full results of mixed linear modeling and post-hoc tests are included in Supplementary Figure 1.

## Discussion

The patients in our cohort who underwent DBS for OCD exhibited striking variability in the timing of benefit from the therapy, with clinical outcomes demonstrating two trajectories in time-to-response. Of the 16 responders in our cohort, we observed a dichotomy in response latency: rapid responders achieved clinically significant response within three months of stimulation initiation, and slow responders required more than three months of continuous stimulation to achieve response. Lead localization revealed variability in electrode placement between these two groups. Rapid responders’ active contacts were more superiorly placed in the right hemisphere relative to those of slow responders and trended toward a more posterior placement, particularly in the left hemisphere. Diffusion tractography revealed that, in the right hemisphere, rapid responders demonstrated robust recruitment of streamlines to the SF cortex early in their treatment course. The modest decline in streamline counts over treatment time may be due to a slight reduction in stimulation amplitude in several patients across the treatment course. In contrast, slow responders showed an increase over time in streamlines to the SF and cOF cortices throughout their treatment course, suggesting that greater and broader PFC recruitment was needed before clinical improvement emerged.

While the course of response to DBS for OCD likely reflects a mix of immediate neuromodulation and slower plasticity-driven remodeling of dysfunctional circuits, our study suggests that broader engagement of particular PFC projections is associated with faster clinical response (Shea et al., 2025). When the active contact is positioned to optimally engage the PFC network, lower amplitudes likely suffice to achieve comparable therapeutic benefits. In contrast, the slow responders required longer-term therapy of on average 294 ± 131 days (range 154-489 days), necessitating progressive stimulation up-titration, presumably to recruit a larger number of fibers targeting clinically significant PFC regions. Importantly, long-term follow-up outcomes did not significantly differ between the two groups. Slow responders eventually reached comparable efficacy, but they required stimulating more fibers to do so. Collectively, these findings support the hypothesis that more effective lead positioning relative to critical white matter fiber populations facilitates faster clinical response, but less effective positioning can be overcome by altering the activated contact and/or increasing amplitudes to engage more white matter fibers, and potentially additional time. Rapid and slow responders may also have different OCD symptomatology profiles, which may interact with their baseline white matter organization and/or ability to respond to DBS therapy.

The various OCD DBS targets lie at different points along the ALIC, a large white matter bundle connecting the PFC with the thalamus, STN, and brainstem. The ALIC is highly topographically organized: projections from dorsal PFC regions, such as the dorsolateral PFC (dlPFC), course dorsally, while ventral PFC fibers, including those from the OFC and ventromedial PFC travel ventrally (Lehman et al., 2011; Safadi et al., 2018). This connectivity was reflected across our cohort as more dorsal and posterior-leaning leads engaged more dorsal PFC components, such as the SF cortex. While this general organization is consistent across population-averaged data (Sretavan et al., 2024) and treatment-resistant psychiatric patients (El Jammal et al., 2026), there is inter-individual variability, with comparisons of voxel-wise representations of individual ALIC anatomy showing ∼66.2 % accuracy, emphasizing the importance of subject-specific tractography (Nanda et al., 2017). Given that distinct ALIC subregions mediate connectivity to functionally diverse PFC regions, subtle variations in lead positioning may influence specific PFC networks implicated in OCD pathology. Our study offers insight into how the use of patient-specific tractography may predict variable symptom responses.

Dysfunction of the OFC and dlPFC, regions strongly engaged across our cohort of responders, is implicated in the pathophysiology of OCD. Functional connectivity studies frequently show OFC hyperconnectivity with the striatum in OCD patients, with evidence that pharmacotherapy helps attenuate this pattern (Beucke et al., 2013). Symptom burden also correlates to OFC-striatal coupling strength, with higher Y-BOCS scores demonstrating a greater degree of connectivity (Harrison et al., 2009). Relative to healthy controls, the OFC in OCD patients has lower neuronal densities (de Oliveira et al., 2019) and reduced expression of genes responsible for encoding proteins relevant to excitatory synaptic signaling (Piantadosi et al., 2021). OFC functional activity is also disrupted during task performance in OCD. For example, the OFC in OCD patients fails to properly activate during reversal learning (Chamberlain et al., 2008). In contrast to findings in the OFC, dlPFC hypoactivity contributes to cognitive inflexibility and executive dysfunction in OCD (Fischer et al., 2016). Abnormalities in brain metabolism (Park et al., 2017) and disrupted local synchronization of spontaneous neural activity have been observed in the dlPFC (Chen et al., 2016). OCD patients have reduced dlPFC volumes compared to controls (Norman et al., 2016). Although certainly not the sole regions involved, the dlPFC and OFC do emerge as particularly relevant to OCD. Indeed, it is possible to frame the disorder as an imbalance between dlPFC-dorsal striatal and OFC-ventral striatal loops (McGovern and Sheth, 2017). Within this framework, simultaneous engagement of both prefrontal pathways may produce more efficient alleviation of OCD symptoms.

Clinical neuromodulation findings also underscore the involvement of both the OFC and dlPFC fibers in mediating therapeutic effects of neuromodulation. Hartmann and colleagues (2015) found that fibers projecting to the dlPFC were more common in responders. They also found that projections to the OFC predominated in non-responders, although the specific OFC regions implicated were more lateral than those in our study (Hartmann et al., 2015). Mosely and colleagues (2021) reported that connectivity between the right lead stimulation volume and right OFC was strongly correlated with Y-BOCS reduction (Mosley et al., 2021). Similarly, analyses of OCD patients treated with STN stimulation revealed that optimal symptom alleviation was linked to connectivity of the STN to the dlPFC and OFC, among other regions (Hollunder et al., 2024). Reductions in OFC hyperactivity are correlated with clinical improvement following VC/VS stimulation (Rauch et al., 2006). In our study, both responder types showed robust OFC engagement; however, dlPFC engagement emerged as a defining feature of rapid response to VC/VS stimulation. Results from non-invasive neuromodulation further support the therapeutic relevance of both prefrontal targets. Repetitive transcranial magnetic stimulation (rTMS) of either the dlPFC or OFC regions have demonstrated efficacy in achieving clinical response (Ruffini et al., 2009; Nauczyciel et al., 2014; Haghighi et al., 2015; Khedr et al., 2022). Indeed, a unified connectomic analysis across many types of neuromodulation treatment for OCD (DBS, rTMS, lesions) identified the OFC and dlPFC as likely involved in most or all forms of effective therapy (along with several other regions) (Baldermann et al., 2021). These studies argue for neuromodulation strategies that recruit both PFC regions as effective for clinical improvement. Although our analyses are not focused on distinguishing effective from ineffective tracts, the speed of response may be related to similar circuitry.

The mechanism by which DBS exerts its therapeutic effect remains debated. Although ‘excitatory’ versus ‘inhibitory’ frameworks persist, converging evidence suggests that high-frequency stimulation acts as a network-level modulator, masking pathological circuits and creating an ‘informational lesion’ that may disrupt the transmission of pathological oscillations (Shea et al., 2025). From this perspective, clinical improvement may not arise from direct activation or suppression of local neurons but from restoration of various dysfunctional networks. Stimulation delivered more posteriorly has a greater opportunity to engage multiple converging white matter tracts. Consistent with this concept, our rapid responders’ active contacts trended toward posterior regions of the ALIC, near the VC/BNST, a region increasingly implicated as a favorable node for targeting a wider representation of PFC circuitry (Karas et al., 2018). Engagement of these posterior fiber populations may thus facilitate more comprehensive and efficacious modulation of both limbic and prefrontal circuitry, facilitating faster clinical response.

We acknowledge limitations of our study. Our sample size was modest (N = 16 responders), limiting generalizability and statistical power. The retrospective design prevented standardized programming protocols which could have introduced bias in stimulation settings. Further, dMRI was not available for all participants, limiting tractography analysis to a subset of patients and potentially leading to underrepresentation of variability. Given that non-responders were excluded, these results do not address the relation between connectivity patterns and treatment failure. Future studies in larger, prospective cohorts with harmonized imaging and programming protocols would help address these limitations.

## Conclusion

This study highlights that response latency is an important but underexplored dimension of DBS outcomes in treatment-refractory OCD. We demonstrate that rapid improvement may be associated with earlier recruitment of dlPFC projections, while delayed response trajectories may reflect progressive engagement of OFC and dlPFC fibers with time and changes to stimulation settings. These findings suggest that structural connectivity between VTAs and different PFC regions influences not only the magnitude but also the timing of therapeutic benefit.

## Data Availability

All data produced in the present study are available upon reasonable request to the authors.

## Acknowledgments

This research was supported by the National Institutes of Health (NIH) National Institute of Mental Health (NIMH) R01MH139889 (N.R.P.), NIH UM1NS32207 (S.R.H.), and NIH BRAIN Initiative via UH3NS100549 (W.K.G.). Additional support was provided by the Robert and Janice McNair Foundation (N.R.P., S.R.H., S.A.S.), the Gordon and Mary Cain Pediatric Neurology Research Foundation Laboratories at Texas Children’s Hospital (S.A.S.), and a NARSAD Young Investigator Grant from the Brain & Behavior Research Foundation (N.R.P.).

